# Assessment Of Anastomotic Leak During the Implementation of Laparoscopic Surgery for Rectal Cancer in Morocco: 21-years Retrospective Study

**DOI:** 10.64898/2026.03.21.26348966

**Authors:** Sarah Benammi, Hamza Sekkat, Youness Bakali, Mouna El Alaoui, Farid Sabbah, Mohamed Raiss, Abdelmalek Hrora

## Abstract

**Background:** Anastomotic leakage (AL) is a major postoperative complication following rectal surgery, leading to increased morbidity, poor oncologic outcomes, and reduced quality of life. While significant data exist regarding rectal cancer management in developed countries, studies from developing nations remain limited. This study aims to evaluate the incidence of AL in rectal cancer surgery during the introduction of minimally invasive surgery (MIS) at a referral center in a developing country.

**Methods:** A retrospective study was conducted at Ibn-Sina University Hospital from 2001 to 2022, including patients who underwent curative rectal cancer surgery. Data were analyzed descriptively with continuous variables presented as mean, median, and standard deviation, while categorical variables were reported as frequencies and proportions. Univariable analysis identified risk factors associated with AL, using appropriate statistical tests for continuous and categorical variables.

**Results:** A total of 306 patients were included, with 39.9% undergoing minimally invasive procedures and 60.1% open surgeries. AL occurred in 16.3% of cases, with higher rates (19.1%) before 2014, compared to 9.1% afterward. Pre-2014, tumor location (p=0.011), surgical resection (R0 vs. R1, p=0.002), and the use of a diverting stoma (p=0.008) were associated with AL. Post-2014, no significant risk factors were identified for AL.

**Discussion:** This study provides valuable insights into rectal cancer surgery outcomes in a developing country. AL rates aligned with global data, showing a reduction after the implementation of MIS. The risk factors identified before 2014 can be attributed to surgical complexity, while the low AL rate post-2014 suggests improved surgical techniques.

**Conclusion:** Assessing the risk of AL is vital for early intervention and optimal surgical planning. The study highlights the advantages and challenges of implementing minimally invasive surgery in developing countries, emphasizing the need for more data in such settings.

## INTRODUCTION

Prevalence of colorectal cancer has been dramatically growing in recent years worldwide. According to World Health Organization, colorectal cancer is the third leading cause of cancer-related deaths, in both male and female patients globally (1). It is the fourth most diagnosed cancer in female patients in Morocco (2). Moreover, while data is reported abundantly in developed country, colorectal cancer statistics from developing countries is scarce, thus requiring further investigations reporting data from developing countries’ perspective and environmental conditions.

Among the most frequent complications of rectal surgery, anastomotic leakage is the most detrimental postoperative complication in terms of the impact on morbidity, mortality and quality of life (3-6). Anastomotic leak significantly increases postoperative morbidity, especially in low anterior resection (LAR) for lower rectal cancer (4, 5). In addition, it impacts the patient’s quality of life significantly (4, 7). AL may impair functional outcome as well including reduced neorectal capacity, evacuation problems, fecal urgency, and incontinence (8, 9). Finally, postoperative complications may delay adjuvant postoperative chemotherapy which could lead to an increased recurrence rate and a poor oncologic prognosis. Alongside these complications and the potential need for reoperation, anastomotic leakage management consumes extensive healthcare resources and expenses (6, 10). More data reflecting on developing countries perspective is therefore essential. Moreover, it is essential that patients should be informed adequately about said complications to allow the most informed consent possible making this assessment vital.

The aim of our study was to study anastomotic leak after rectal cancer surgery during the period of minimally invasive surgery implementation from a developing country perspective in a tertiary university hospital center.

## MATERIALS AND METHOD

This was a retrospective study based on patient’s charts review from 2001 to 2022. The study was conducted at the surgical department « C » in the University Hospital Ibn-Sina in Rabat, Morocco. All patients admitted for rectal cancer treatment were included. No age limit was implemented. All patients who underwent surgical management were included, both curative and palliative surgeries. We excluded patients admitted for rectal cancer management who didn’t undergo surgery.

We collected socio-demographic and anthropometric data using patient’s chart review (Age of participants; Gender (Male or female); ASA score; Preoperative hemoglobin; Preoperative Albuminemia). We included preoperative disease characteristics (Biological marker ACE (carcinoembryonic antigen); Biological marker CA19-9 (cancer antigen); Tumor location; Neo-adjuvant therapy: including all applicable protocols (chemotherapy, chemo-radiotherapy). We collected procedural characteristics (Surgical approach: laparoscopic approach either total and partial, open surgery, or non-applicable; Surgical indication: curative R0 vs palliative R1; Ostomy creation; Survival status; Reported on charts disease recurrence).

Postoperative outcomes were divided into clinical outcomes (Postoperative complications <90days according to Clavien-Dindo classification; Anastomotic leak: reported in charts diagnosed both clinically and or with para-clinical means.); and disease management outcome (Positive resection margins on surgical specimen histology assessment; Total number of collected lymph nodes on surgical specimen histology assessment; Total number of positive lymph nodes on surgical specimen histology assessment; Positive vascular invasion on surgical specimen histology assessment; Positive nerve invasion on surgical specimen histology assessment; Adjuvant therapy).

We defined anastomotic leak as defect in the intestinal wall at the location of anastomosis leading to a communication between the intra- and extraluminal compartments. Diagnosis was suspected in instances of positive CRP on postoperative day 2 and day 4 and confirmed upon CT-scan confirming parietal defect or intraperitoneal fluid collection surrounding the anastomosis. The overall incidence of anastomotic leak (AL) and overall postoperative complications according to Clavien-Dindo classification was identified, and the incidence of AL was further summarized by tumor location (low rectum, mid-rectum, high rectum) and surgical approach (open or laparoscopic). Finally, we divided our cohort into two groups: from 2001 to 2023 with more predominant open surgery, and 2014 to 2022 with laparoscopic surgery more predominant.

All study variables were analyzed. Continuous variables were presented as mean, median, and standard deviation, while categorical variables were expressed as frequencies and proportions. Univariable analysis was conducted comparing all variables included in our study to the incidence of AL to define the factors associated with AL. Student’s t-test or Wilcoxon’s rank sum test was applied for comparison of continuous variables while a chi-squared test or Fisher’s exact test was conducted for categorical variables as appropriate after testing assumptions including normality. A p-value of 0.05 or less was considered to indicate statistically significant difference.

The univariable and multivariable logistic regression models were fitted to assess the risk factors associated with AL so that the odds ratio of each risk factor could be calculated. The multivariable model included factors that showed the statistical significance based on univariable analysis. All analyses were conducted using JAMOVI (11). Institute’s ethics committee approval was submitted. Human Ethics and Consent to Participate declarations was not applicable as this was a retrospective study based on patient’s chart review. There was no Funding for this study, and no clinical trial number applicable.

## RESULTS

A total of 306 patients were included from 2001 to 2022. The mean age ±SD of the study population was 54.8 ± 14.2 years. Female patients represented 50.3% (n=154). All collected variables were detailed in the descriptive analysis (Table 1). Minimally invasive procedures represented 39.9% of all cases, while open surgeries comprised 60.1%. The first case of minimally invasive surgery was recorded in 2005. Minimally invasive surgery was conducted more than open surgery per year starting 2014 (Table 1). AL was reported in 19.1% of cases prior to 2014 vs to 9.1% after 2014. Rectal cancer was 64.3% a low-rectum, followed by mid-rectum with 21.3% (n=64), and finally high-rectum with 14.3% (n=43). Neo-adjuvant therapy was received in 42.2% (n=128) patients. Surgery was curative in 74.6% (n=226).

**Table 1.** Descriptive analysis.

| *Variable* | *% (n)* | *Mean±SD* | *Median*<br>*[min,max]* |
| --- | --- | --- | --- |
| **Age** | (n=306) | 54.8±**14.2** | 57.0 [18, 86] |
| **Gender** |  |  |  |
| *Female* | 50.3% (n=154) |  |  |
| *Male* | 49.7% (n=152) |  |  |
| **Preoperative Hb (g/l)** | (n=266) | 11.8±**1.9** | 12.0 [6, 15] |
| **Preoperative Albuminemia (g/l)** | (n=138) | 36.9 ±**4.5** | 37.0 [24, 50] |
| **ASA** |  |  |  |
| *1* | 39.8% (n=88) |  |  |
| *2* | 6.8% (n=15) |  |  |
| *3* | 0.9% (n=2) |  |  |
| **ACE** | (n=178) | 43.2±**217** | 2.9 [0.3, 21300] |
| **CA 19-9** | (n=110) | **23.9±64.1** | 10.3 [1, 596] |
| **Rectal cancer location** |  |  |  |
| *Low-rectum* | 64.3% (n=193) |  |  |
| *Mid-rectum* | 21.3% (n=64) |  |  |
| *High-rectum* | 14.3% (n=43) |  |  |
| **Neoadjuvant therapy** |  |  |  |
| *Yes* | 42.2% (n=128) |  |  |
| *No* | 57.8% (n=175) |  |  |
| **Surgical approach** |  |  |  |
| *Laparoscopic* | 40.1% (n=120) |  |  |
| *Open surgery* | 40.2% (n=138) |  |  |
| *Non applicable* | 13.7% (n=41) |  |  |
| **Surgical indication** |  |  |  |
| *R0 Curative* | 74.6% (n=226) |  |  |
| *R1 Palliative* | 25.4% (n=77) |  |  |
| **<90days postoperative complications** |  |  |  |
| **Clavien-Dindo** |  |  |  |
| *0* | 65.2% (n=199) |  |  |
| *1* | 8.5% (n=26) |  |  |
| *2* | 12.2% (n=37) |  |  |
| *3* | 0.7% (n=2) |  |  |
| *4* | 9.5% (n=29) |  |  |
| *5* | 39.9% (n=12) |  |  |
| **Anastomotic leak** |  |  |  |
| *Yes* | 16.3% (n=45) |  |  |
| *No* | 83.7% (n=231) |  |  |
| **Positive resection margins** |  |  |  |
| *Yes* | 15.7% (n=41) |  |  |
| *No* | 84.3% (n=220) |  |  |
| **Vascular invasion** |  |  |  |
| *Yes* | 7.2% (n=22) |  |  |
| *No* | 92.8% (n=284) |  |  |
| **Nerve invasion** |  |  |  |
| *Yes* | 10.5% (n=32) |  |  |
| *No* | 89.5% (n=274) |  |  |
| **Adjuvant therapy** |  |  |  |
| *Yes* | 47.1% (n=144) |  |  |
| *No* | 52.9% (n=162) |  |  |
| **Disease recurrence** |  |  |  |
| *Yes* | 27.9% (n=65) |  |  |
| *No* | 72.1% (n=168) |  |  |

**Table 2.**
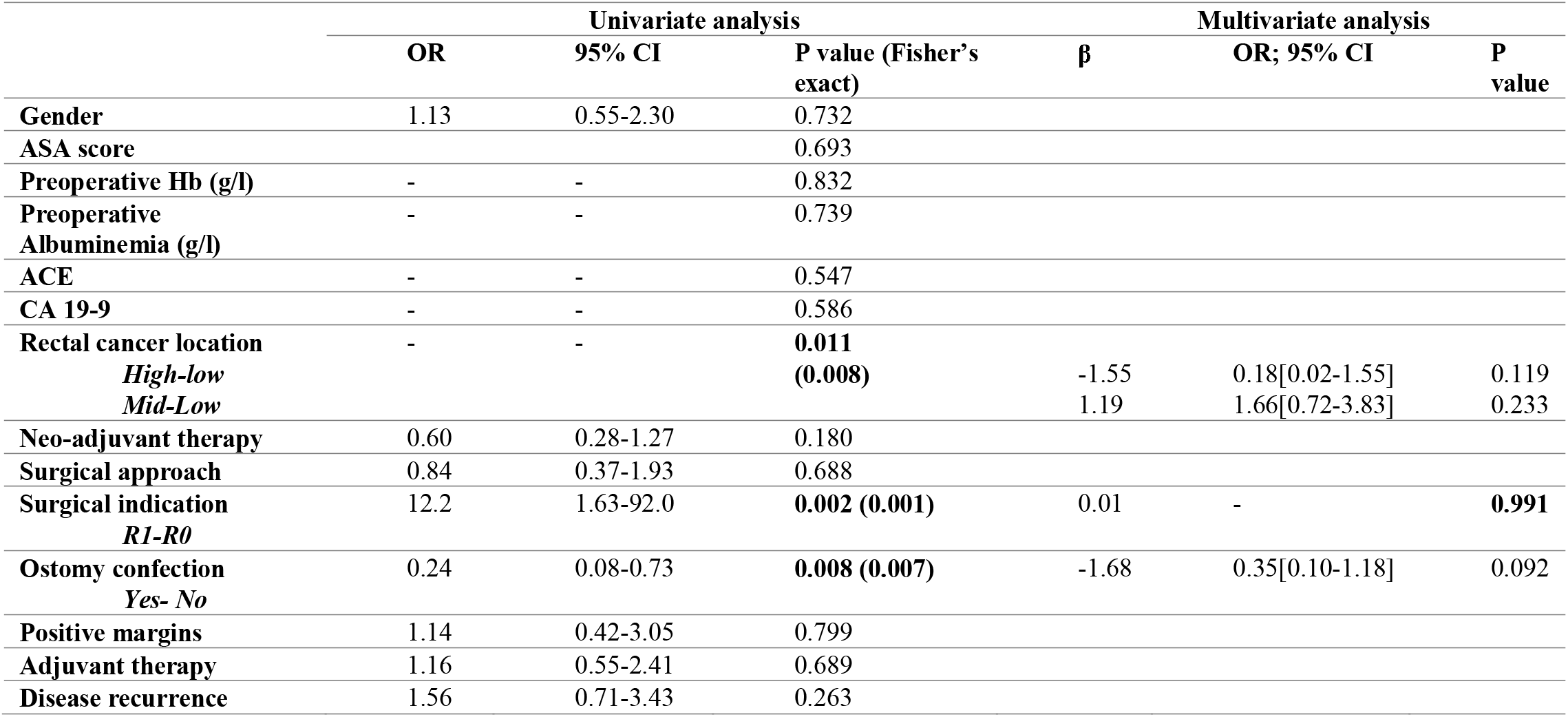
Univariate analysis and Multivariate analysis of anastomotic leak before implementation of minimally invasive surgery from 2001 to 2013)

**Table 3.**
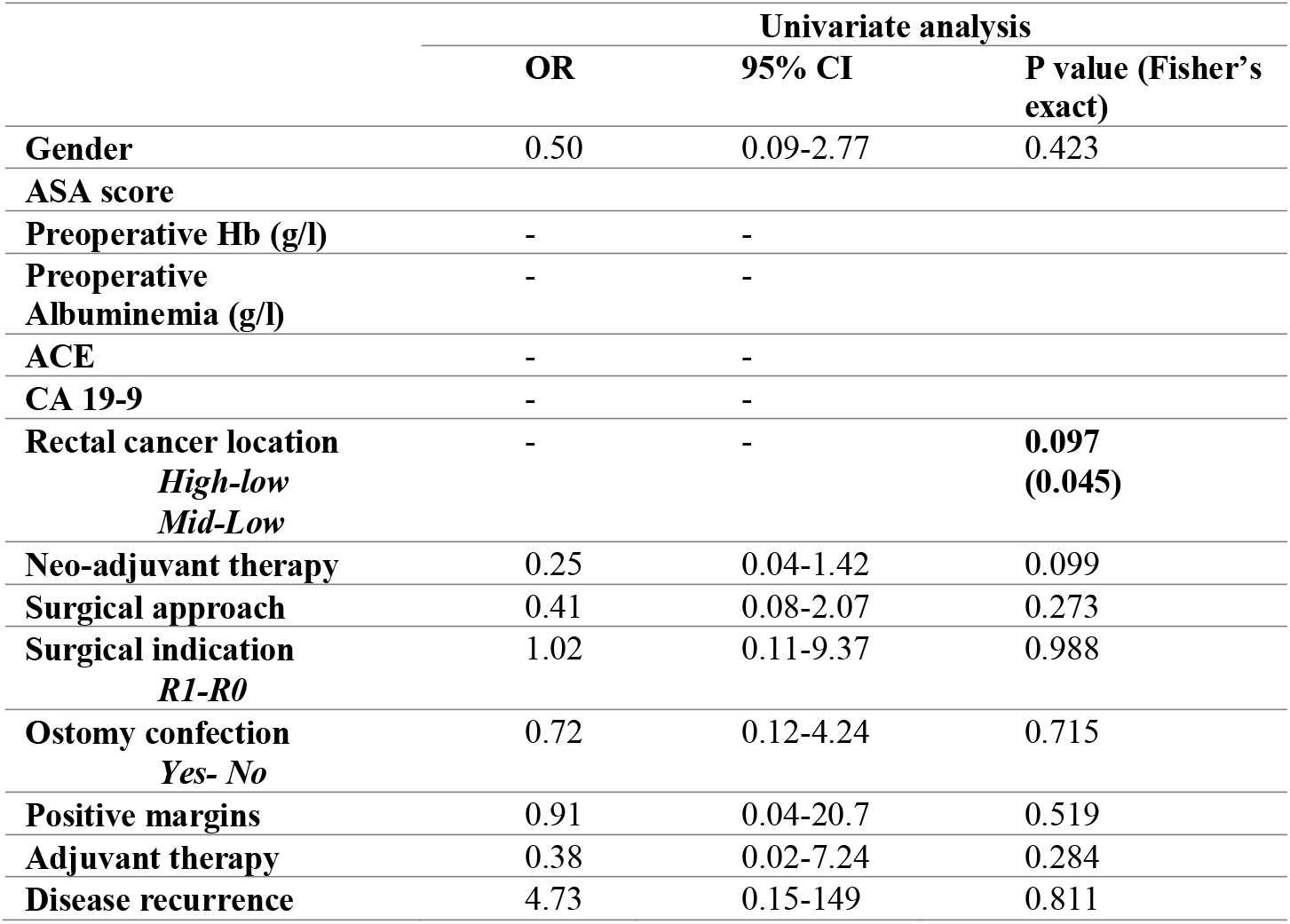
Univariate analysis and Multivariate analysis of anastomotic leak after implementation of minimally invasive surgery (from 2014-2022)

Univariate analysis of the cohort from 2001 to 2013 found that tumor location (p = 0.011), surgical resection (R0 vs. R1) (p = 0.002) and the use of a diverting stoma (p = 0.008) to be associated with AL. Neoadjuvant radiotherapy was not associated with AL (p=0.180). Multivariate analysis found no associated variables to AL. Univariate analysis of the cohort from 2014 to 2022 found tumor location significantly associated with AL on Fisher’s exact text (p=0.045); previous variables were not associated (resection (R0vsR1) p=0.988; use of diverting stoma p=0.715).

## DISCUSSION

Anastomotic leakage is the major postoperative complication in rectal cancer surgery representing the most detrimental among all reported factors in terms of the impact on morbidity, mortality and quality of life (4-6). Globally including all colorectal and coloanal surgeries, the rate of anastomotic leak is ranging between 5% to 36.3%, reportedly depending upon perioperative factors (12-14). In a nationwide published study of the Italian Society of Surgical Oncology, Degiuli et al published an overall anastomotic leak rate after restorative resection for rectal cancer of 10.2% (15). The incidence of anastomotic leakage after low anterior resection varies 3–27% (4, 16). In fact, the wide range of incidence can be explained by the different possible definitions of anastomotic leak. In a review published in 2001 more than 50 different definitions of AL were reported (17). The most common definition was ‘a communication between the intraluminal and extraluminal compartments owing to a defect of the integrity of the intestinal wall at the anastomosis between the colon and rectum or the colon and anus’ (17, 18). Anastomotic leak can be diagnosed upon digital examination, anoscope findings, and radiology testing including enema imaging with an iodinated contrast agent or CT-scan (4). Usually, AL is diagnosed between postoperative days 1 and 14, however, AL can be reported later after patient discharge (14, 15).

Anastomotic leak significantly increases postoperative morbidity, especially low anterior resection (LAR) for lower rectal cancer (4, 5). It has been proven that AL has a short-term postoperative negative impact including quality-of-life detriments, longer hospitalization and cost, and functional impairment such as evacuation problems, fecal urgency, and incontinence (4, 7-9). Additionally, it might necessitate more surgeries to establish either a permanent or temporary stoma (19). Most of all, these complications may delay adjuvant postoperative chemotherapy which could lead to a direct impact on the oncological outcome, an increased recurrence rate and a poor prognosis (19). Such impairment over oncologic prognosis emphasizes the need to further assess risk factors for AL, furthermore according to data specific to local conditions. Regarding long-term outcome effects of AL, multiple studies have been reported to both confirm or dismiss AL as risk factor in oncological prognosis (19). However, the first metanalysis in 2011 observed a significant association between AL and local recurrence (LR), distant recurrence (DR), and cancer-specific mortality (19, 20). Since then, multiple subsequent meta-analysis confirmed these findings (20).

All taken together, risk assessment of AL is crucial for early decision-making. Various risk factors have been described to affect AL and postoperative outcome divided into preoperative and intraoperative due to the high number. Reported preoperative factors are male sex, high BMI, smoking habit, alcohol intake, previous abdominal surgery, prolonged corticosteroid intake (4, 15, 19, 21, 22). Intraoperative factors were related to the surgical variables and tumor characteristics including locally advanced tumor, lymph-node metastases, tumor location, tumor size, minimally invasive approach, a longer operating time, the number of staples used, preoperative chemotherapy, operative duration, blood loss volume and transfusion, left colic artery ligation, and a combined multivisceral resection were all identified as preoperative independent risk factors (4, 15, 19, 21, 22).

Male gender can be related to increased AL risk, probably due to the narrower male pelvis rendering more difficult surgery both open and laparoscopic, as well as androgens that may affect the bowel microcirculation acting on intestinal endothelial function (15, 23). An experimental study on rats showed a less favorable collagen metabolism in colonic anastomoses of males compared with females during early wound healing (24). However, we didn’t find gender to be an associated factor in our study. Greater BMI is a modifiable independent risk factor for AL (15). Goulart et al. showed a direct relationship between visceral fat and anastomotic leakage and reoperation (25). Hence weight and nutritional status are major factors to be considered during colorectal preoperative evaluation (23).

Preoperative albumin has been associated in the past to AL (23, 26). However recent studies have been refuted this theory (27). In our study, we found no significant association between albumin level and anastomotic leakage either in univariate or multivariate analysis.

While some authors showed a statistically significant relationship between preoperative chemotherapy and AL, several others did not confirm thus results (15, 28, 29). In fact, there are many controversial papers regarding the role of neoadjuvant radiotherapy on AL incidence. Arezzo et al. conducted a meta-analysis on transabdominal resection for rectal cancer that reported data about anastomotic leak. They studied the effect of both short- and long-course radiotherapy on AL separately, significant association with short-course treatment was retained (14). Chemoradiotherapy was considered a risk factor for AL in some retrospective studies but was not confirmed in several others (14). However, overall neoadjuvant therapy was not found to be associated with AL (14).

Previous studies have established that the risk of AL is higher in low rectal cancers due to the technical difficulties associated with performing anastomosis in this area, especially when rectal mobilization is inadequate (Rullier). Tumor distance from the anal verge have been identified as a significant factor since 1990s, with Rullier et al. publication reporting a 6.5-times increased risk in anastomoses located <5 cm from the anal verge. More recently Zhang W et al. showed that anastomosis less than 7 cm from the anal verge is an independent risk factor for leakage (30). In fact, though only retained on univariate analysis our study found that rectal cancer distance from the anal verge was significantly associated to AL represented by tumor location. Additionally, it is well-documented that the use of a diverting stoma can mitigate the consequences of AL by preventing fecal contamination of the anastomosis site (31). Although our study found a significant association between the use of a diverting stoma and AL in the earlier cohort, no such association was observed in the more recent years, possibly due to the improved technique and outcomes associated with MIS.

As evidenced by our data, AL rate dropped from 19.1% to 9.1% after implementation of more predominant minimally invasive surgery after 2014. This reduction could reflect improvements in surgical technique and the potential benefits of MIS, including better visualization, precision, and faster recovery times (32). However, our univariate analysis indicated that tumor location, surgical resection margin (R0 vs. R1), and postoperative complications as classified by the Clavien-Dindo system were significantly associated with AL in the cohort prior to 2014 in contrast to post-2024 cohort showing no statistically significant associations between these variables and AL. This could indicate that the factors influencing AL are multifactorial and may vary depending on the surgical approach. Further research is essential to identify modifiable risk factors and optimize management strategies

However, this study is subject to several limitations. Being retrospective in design, it relies heavily on the accuracy and completeness of patient chart data. Additionally, the disparity of multivariate analysis findings between the two cohorts prior and post-2014 can be due to the low number of positive AL. Further multicentric research is needed to evaluate minimally invasive rectal surgery outcome and risk factors that may contribute to AL and other postoperative complications for generalizable results.

## CONSLUSION

In conclusion, anastomotic leakage remains a significant complication following rectal cancer surgery, with substantial impacts on morbidity, survival, healthcare resources and oncological outcome making the management of AL a priority in rectal cancer surgery. Moreover, given the rising burden of colorectal cancer in developing countries, this study contributes to literature regarding rectal cancer surgery data, considering the importance of addressing these complications within local contexts to improve patient outcomes and healthcare delivery.

The implementation of minimally invasive surgery may have contributed to a reduction in AL rates in our cohort. However, AL risk factors are numerous and are multifactorial. Therefore, further research is essential to identify modifiable risk factors and optimize management strategies

Our results provide insights into the surgical outcomes, particularly the rate of AL, which remains a significant postoperative complication with considerable implications for both patient outcomes and healthcare resources. This study highlights that while the incidence of colorectal cancer is on the rise in Morocco, nonetheless there is still a gap in comprehensive reporting and follow-up, which is necessary for developing region-specific guidelines.

### The Human Ethics and Consent to Participate declarations are not applicable as this retrospective chart review study

Involved patients were informed and consent was obtained at admission about inclusion in clinical research at our university hospital, and multimedia use in compliance with anonymity protections.

**There was no Funding for this study No clinical trial number applicable**.

## Data Availability

All data produced in the present study are available upon reasonable request to the authors

## Notes

**Conflict of interest:** None to declare.

**Competing interest:** We have no conflict of interest to report.

### Competing Interest Statement

The authors have declared no competing interest.

### Funding Statement

This study did not receive any funding

### Author Declarations

Ethics committee IRB of Faculte de Medecine et de Pharmacie de Rabat gave ethical approval for this work

